# Signal properties and stability of a chronically implanted endovascular brain computer interface

**DOI:** 10.1101/2025.09.19.25335897

**Authors:** Nikole Chetty, Kriti Kacker, Ariel K Feldman, Peter E Yoo, James Bennett, Adam Fry, Idan Tal, Nicholas F Hardy, Sadegh Ebrahimi, Cesar Echavarria, Abbey Sawyer, Hunter R Schone, Noam Y Harel, Raul G Nogueira, Shahram Majidi, Elad I Levy, Amit Kandel, Katharine (Katya) Hill, Nicholas L Opie, David Lacomis, Jennifer L Collinger, Thomas J Oxley, David F Putrino, Douglas J Weber

## Abstract

**Background:** Implanted brain-computer interfaces (iBCIs) establish direct communication with the brain and hold the potential to enable people with severe disability to achieve control of digital devices, enabling communication and digital activities of daily living. The ability to access brain signals reliably and continuously over many years post-implantation is crucial for iBCIs to be effective and feasible. This study investigates the signal characteristics and long-term stability of neural activity recorded with a stent-electrode array over 1 year post-implant.

**Methods:** We report on five participants with paralysis who were enrolled in an early feasibility clinical trial of an endovascular iBCI (Stentrode; ClinicalTrials.gov, NCT05035823). Each participant was implanted with a 16-channel stent-electrode array, deployed in the superior sagittal sinus to record bilaterally from the primary motor cortices. Neural activity was recorded during home-based sessions while the participants performed a set of standardized tasks. Metrics including motor signal strength during attempted movement, resting state signal features, and electrode impedances were quantified over time.

**Results:** Motor-related modulation in neural activity was exhibited in the high-frequency bands (30-200 Hz) during attempted movements, with rest and attempted movement states showing sustained differentiation over time. Impedance and resting state band power for most channels did not change significantly over time.

**Conclusions:** These findings provide strong evidence that the endovascular BCIs may be suitable for long-term neural signal acquisition in the home environment, demonstrating the ability to record movement-related modulation over one year.

## Introduction

Implanted brain-computer interfaces (iBCIs) hold great promise for restoring communication and digital device control for people with profound paralysis due to trauma or disease^1^. By measuring electrical activity generated by neurons in motor areas of the brain, especially the motor cortex, BCIs form a conduit for expressing motor intentions, goals, and needs^2^. For people with severe disability, an iBCI may serve as the only option for personal expression, enabling control of a cursor for spelling^3^ and even communication via artificial speech synthesis^4^.

iBCI systems that are under development use a variety of interface technologies to measure neural activity in the brain, with each approach and implantation site offering trade-offs including signal fidelity, spatial resolution, long-term stability, and surgical risk. Intracortical microelectrode arrays (MEAs) offer the unique ability to detect action potentials from individual neurons, as well as local field potentials. However, intracortical MEAs require a craniotomy and dural resection, which pose a small, but non-zero risk of infection and tissue damage and may be susceptible to material degradation^5,6^. Furthermore, spike-based iBCIs typically require frequent recalibration or decoder updates, such as manifold alignment, to overcome signal instability^7^. Nonetheless, MEAs give access to rich movement-related information and have enabled several notable human iBCI milestones, including computer control, robotic arm control, and speech decoding^8–11^.

Electrocorticography (ECoG) is another iBCI technology that uses arrays of non-penetrating microelectrodes placed above or below the dura to record field potentials. ECoG-based iBCIs typically utilize spectral features from the sensorimotor cortex during imagined or overt movement, including beta de-synchronization and, most notably, gamma synchronization which provides rich, focal information^12–15^. ECoG-based iBCI studies have demonstrated successful computer cursor control, speech decoding, and digital communication in people who are locked-in^16–19^. Nevertheless, the placement of ECoG electrodes necessitates a craniotomy, burr holes or slits, limiting its use to specific clinical scenarios.

Endovascular electrode arrays are a newer type of iBCI recording array that can be delivered via catheter introduced in the jugular vein, thereby offering a minimally invasive alternative to the aforementioned iBCIs. Endovascular electrode arrays can be implanted in the cerebral venous system adjacent to cortical targets, e.g. the superior sagittal sinus adjacent to the medial wall of primary motor cortex of both hemispheres of the brain. This approach enables neural signal recording from within the blood vessel wall without directly penetrating brain tissue. A first-in-human clinical trial of stent-electrode array based iBCI demonstrated safe implantation and successful decoding in four individuals with severe bilateral upper-limb paralysis^20,21^. However, given that endovascular electrode arrays were developed relatively recently, there are limited reports on the spectral features and long-term signal quality of these vascular ECoG (vECoG) signals. For any iBCI to be viable for long-term use, signals must remain stable over time to enable decoding of user intent. Multiple factors could contribute to signal instability and/or loss of iBCI functionality in people with amyotrophic lateral sclerosis (ALS), including neuronal degeneration and cortical atrophy, cognitive decline, biological response, and device-related failures. Here, we report on the signal properties and long-term stability of vECoG recordings from five participants implanted with a stent-electrode array as a part of the COMMAND Early Feasibility Study (NCT05035823).

## Results

Six participants underwent implantation of a stent-electrode array (Stentrode), which was delivered endovascularly via catheter to a target location in the superior sagittal sinus adjacent to motor cortex (Fig. 1, Table 1). To investigate the stability of these signals over time, we analyzed three key metrics: motor signal strength during attempted movement, resting state signal features, and electrode impedances across the duration of each participant’s involvement in the study, up to 12 months. The data presented here comprises a subset of the total dataset acquired during the early feasibility study and was analyzed independently in an exploratory analysis to evaluate signal properties and stability. A separate manuscript will report clinical outcomes and safety data.

**Fig. 1:**
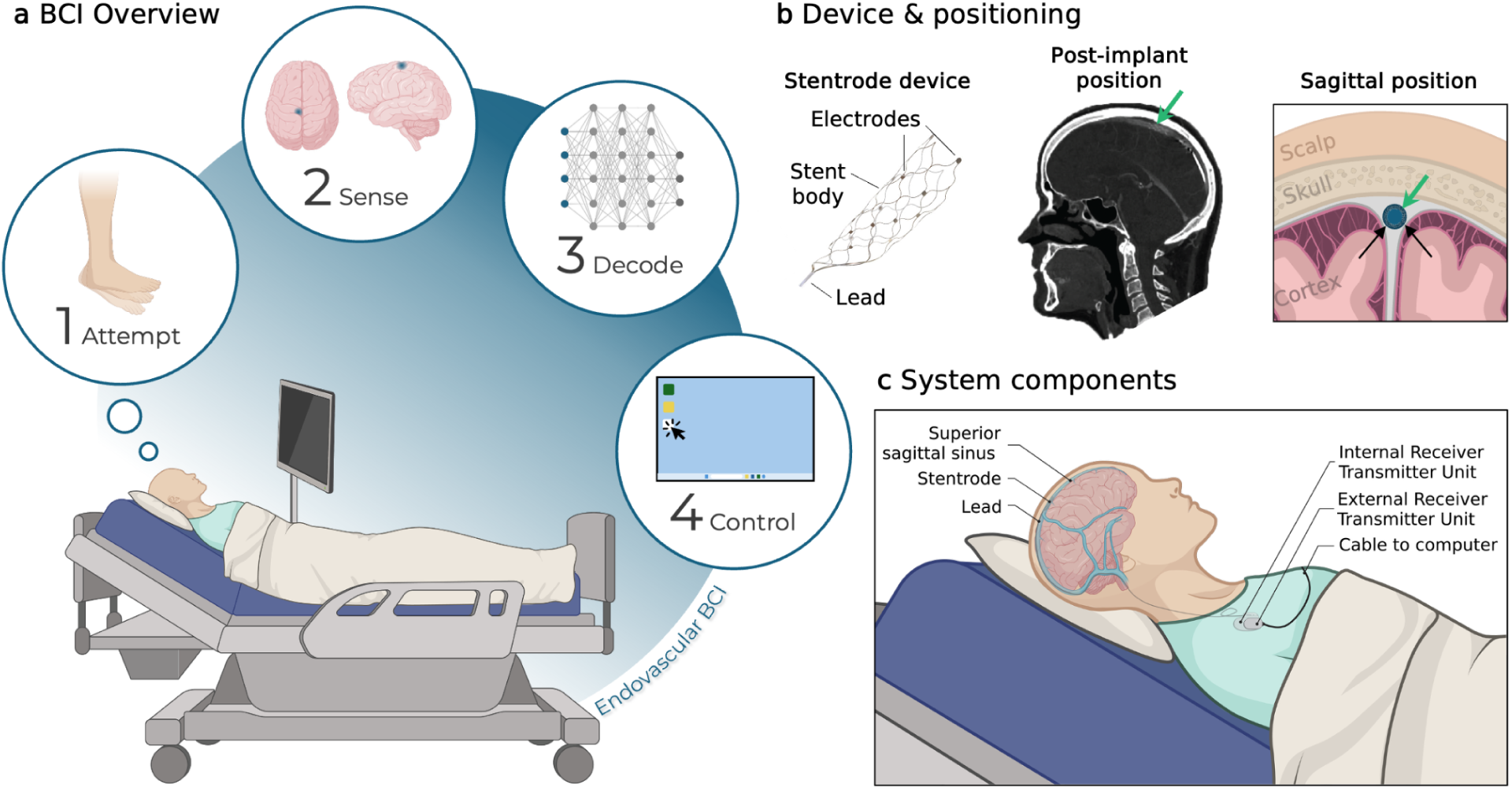
BCI setup and device location. **a**, Schematic of the click-based brain-computer interface (BCI). Neural activity of the participant attempting to move was recorded from the implanted stent-electrode array and transmitted to a computer for decoding and computer control. **b**, Left: Stent-electrode array with 16 recording electrodes embedded on a stent. Middle: Example CT showing the post-implant location of the stent-electrode array (green arrow) in the superior sagittal sinus adjacent to motor cortex. Right: Sagittal plane showing the stent-electrode array (green arrow) is deployed in the superior sagittal sinus (blue), adjacent to motor cortex. **c**, Components of the fully implanted BCI system. The lead connects to the internal receiver transmitter unit (IRTU) in the subcutaneous, subclavicular pocket. The IRTU communicates with an external receiver telemetry unit (ERTU), which relays the signals to a signal control unit and then a laptop. Parts of this figure were created with BioRender.com.

**Table 1.** Participant demographics.

| Participant ID | Gender | Age Range (years) | Diagnosis | Time since diagnosis (years) | Clinical Manifestations | Data Follow-up (months) | Data Analysis Scope |
| --- | --- | --- | --- | --- | --- | --- | --- |
| 1 | M | 66 - 70 | ALS | 11 | Severe paralysis, tracheostomy, ventilator dependent, anarthria | 6 | Full |
| 2 | M | 71 - 75 | ALS | 5 | Severe paralysis, tracheostomy, ventilator dependent, anarthria | 12 | Full |
| 3 | M | 51 - 55 | ALS | 3 | Severe paralysis, tracheostomy, ventilator dependent, anarthria | 6 | Partial - Resting state & impedance analysis only |
| 5 | F | 51 - 55 | ALS | 6 | Paralysis, supplemental oxygen, anarthria | 12 | Full |
| 6 | M | 61 - 65 | ALS | 3 | Upper limb paralysis, ambulatory, normal speech | 12 | Full |

ALS: Amyotrophic lateral sclerosis

### Attempted movement modulates endovascular neural signals

An example single-channel broadband and band-limited signal, along with the corresponding spectrogram and PSD (across all trials), is shown during attempted movement of both ankles (Fig. 2a). Following the movement cue, synchronization is observed in the low and high gamma bands compared to rest. As expected and consistent with other recording modalities, the power spectrum follows a 1/f relationship, with power decreasing as frequency increases.

**Fig. 2:**
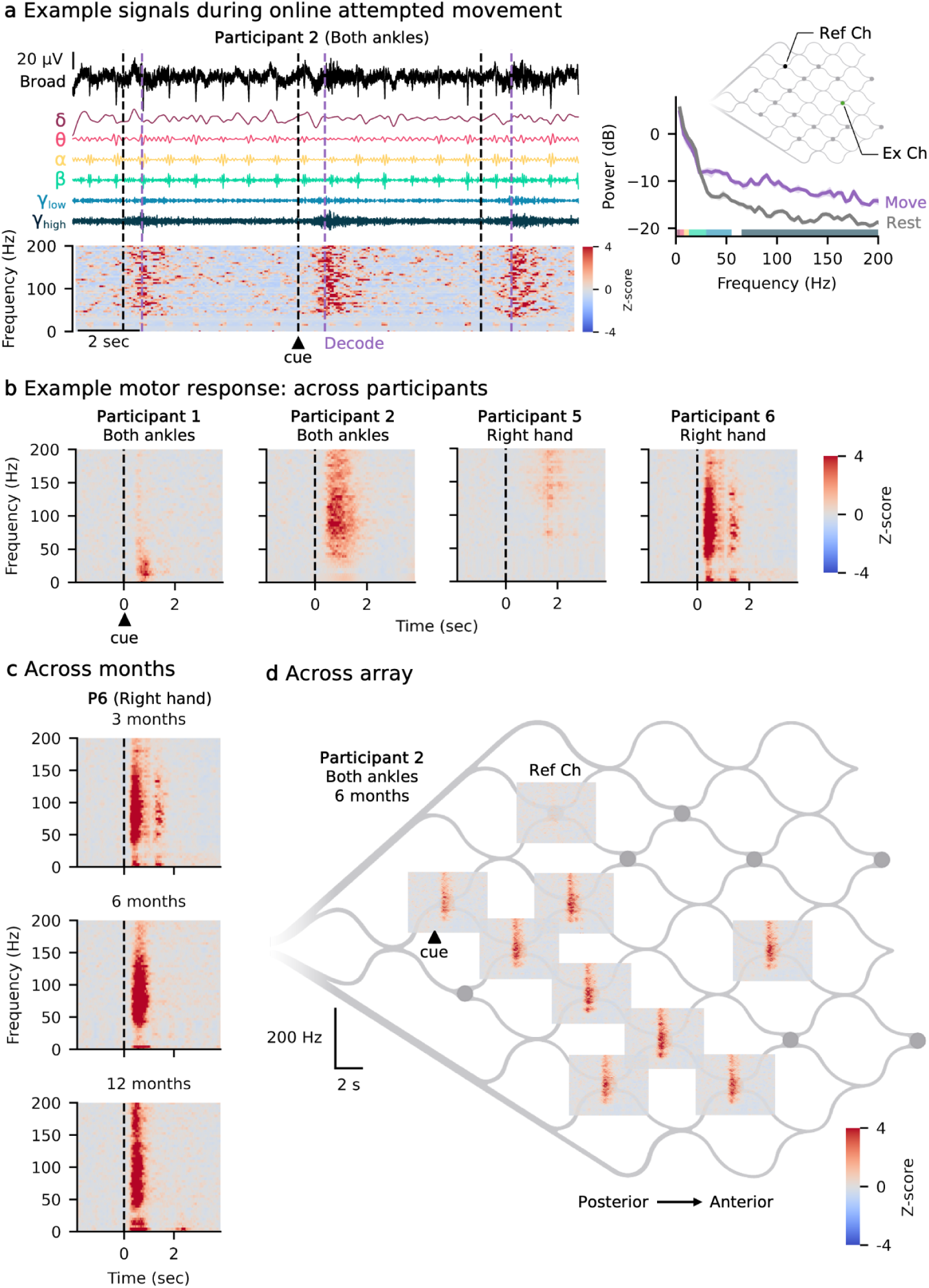
Signal modulation during attempted movement. **a**, The broadband signal from one electrode (top) is shown alongside its corresponding band-limited signals (middle) and spectrogram (bottom) while the participant is being cued to move. The black dashed line indicates the time when the participant was visually cued to move, while the gray dashed line marks when movement was decoded. Right: Power spectral density (PSD) comparing move and rest conditions, highlighting frequency-specific differences in neural activity, especially in the low gamma and high gamma ranges. Solid line represents the mean across trials; shaded regions show the 95% confidence interval of the mean. **b**, Example channel spectrograms at 3-months post-implant for the four participants, showing the trial-averaged time-frequency responses following the visual move cue (indicated by the black dashed line). Red indicates synchronization from rest, while blue indicates de-synchronization. **c**, Spectrograms from the same channel across three time points (3, 6, and 12 months) while the participant attempted right hand movement following the visual move cue (indicated by the black dashed line), demonstrating the similarity of the control signal over time. **d**, Spectrograms from all active channels across the array.

Figure 2 illustrates examples of attempted movement modulation, with formal quantification of modulation strength provided in subsequent analyses. Mean spectrograms from a representative channel from each participant illustrate the variability in signal modulation following the move cue (Fig. 2b). The spectrograms show the mean across 30 trials of attempted movement, aligned to the go cue and z-scored relative to the rest period of each frequency bin (2 seconds preceding the go cue). Mean spectrograms from a single channel over time demonstrate the consistency of spectral responses over the course of the study (Fig. 2c). Mean spectrograms across the array show similar time-frequency modulation on multiple channels during attempted movement of both ankles for P2 (Fig. 2d).

Each participant was tested on their ability to use the BCI to generate binary command signals, akin to a mouse-click. Participants asserted a click command by attempting to move their ankles or hands; the specific gesture for each participant was selected based on a combination of neural modulation feedback, participant preference, and intuitiveness. For P1, the primary gesture was both ankles, which consisted of attempted plantar flexion of the ankle, described as pressing down on a gas pedal. For P5 and P6, the primary gesture was right hand (finger flexion of all digits on the right hand), described as attempting to make a fist. P2 used both ankles for the first 8 months and then switched to the right hand.

### Modulation during attempt movement is stable over time

To evaluate motor signal strength, we measured the Euclidean distance between ‘rest’ and ‘move’ band power in the beta, gamma, and high gamma frequency bands across all channels. This metric quantifies the separation between rest and movement-related neural states, providing insight into the amplitude and stability of task-related neural signal modulation over time. Signals were band-limited, the envelope extracted and smoothed with a 100 ms window, and RMS amplitudes for each channel were computed within rest and movement epochs. Fig. 3a shows examples of the high-gamma motor signal modulation for two channels. The cross-validated Euclidean distance measures the distance between the centroids of the clusters formed in the N-dimensional space formed by the N number of active channels in the stent-electrode array. The mean distance across participants for the beta, low gamma, and high gamma bands was 5.6, 5.8, and 10.9 standard deviations from rest (Fig. 3b). Across all participants, the percentage of sessions exhibiting statistically significant Euclidean distance was 40.33 ± 39.32%, 66.51 ± 22.53%, and 89.32 ± 15.28% for the beta, low gamma, and high gamma bands, respectively. Many factors may contribute to the small magnitude of some observed values. First, the band power metric is not derived directly from the features utilized for decoding, meaning that participants may not be optimizing or learning to modulate these specific features (particularly the beta band). Second, the signal often peaked after the decode window (Fig. 3a), underscoring that this metric captures the distance at the time of decoding rather than at the peak neural response. Third, participants may adopt a strategy of ‘greedily minimizing effort’,^22^ where they adjust their movements to use the minimum effort possible to produce a decoded click. Consistent with this, P6 exhibited a significant decline in high gamma modulation without a corresponding decrease in online accuracy, likely because he learned that such high levels of “effort” were not necessary to elicit a click. Although this adaptive behavior and plateau may be better captured by an exponential decay, a linear fit was utilized in all cases for simplicity and consistency.

**Fig. 3:**
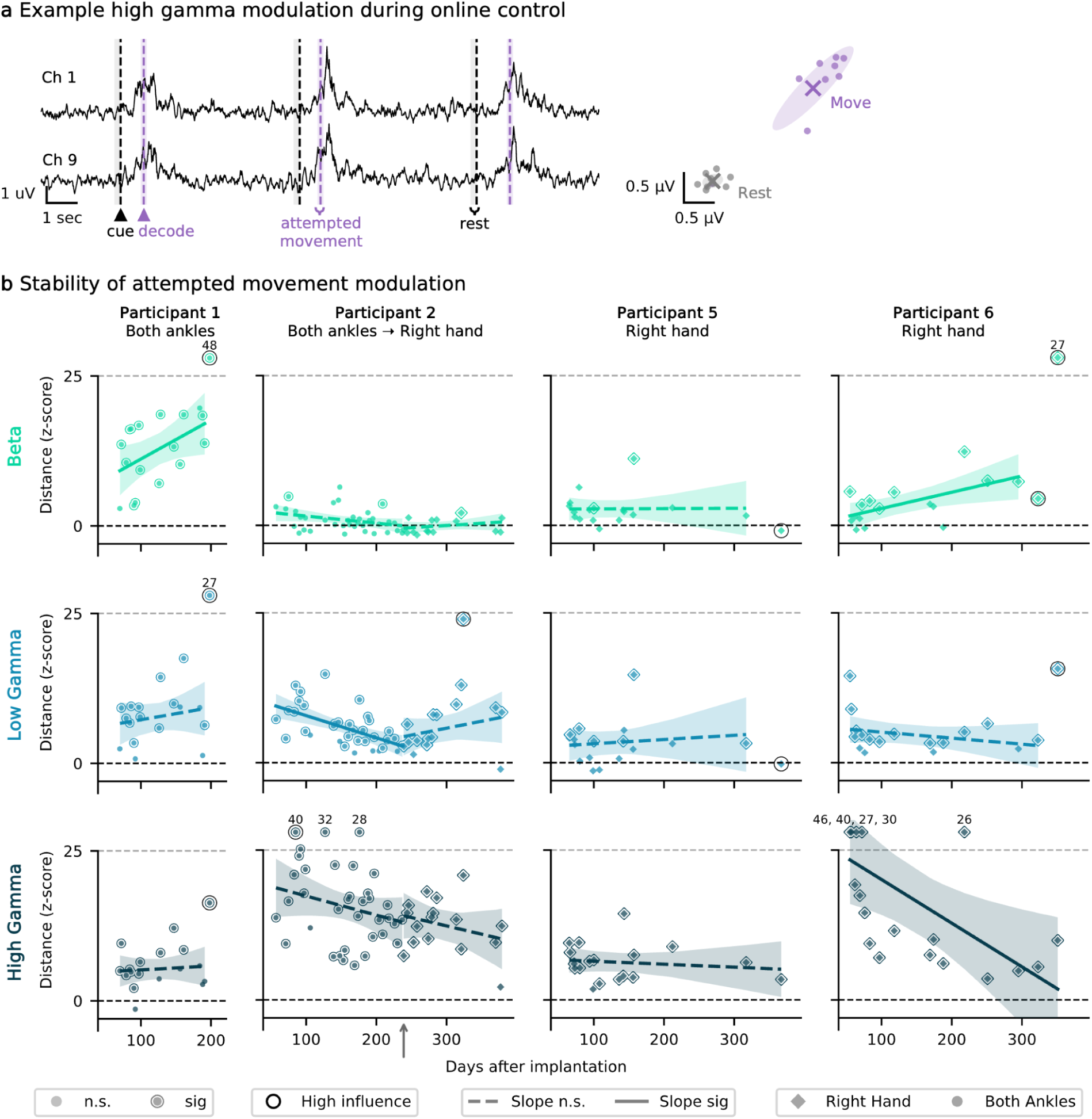
Amplitude and stability of motor signal modulation over time. **a**, Example smoothed envelopes for two channels of high gamma modulation during the command control task (left) (shown for Participant 2 with attempted movement of both ankles at 3 months post-implant). The shaded regions indicate the epochs used to compute the RMS for both attempted movement (purple) and rest (gray). On the right is an example 2D visualization of the move and rest clusters, formed from the two example channels on the left, from which Euclidean distance was calculated. Each scatter point is the RMS of the envelope from a single rest or move attempt. Shaded ellipse shows the covariance and X marks the centroid. The cross-validated Euclidean distance is computed as the distance between the rest and move condition vectors formed in an N-dimensional space, where N number of active channels in the stent-electrode array. **b**, Cross-validated Euclidean distance across participant and frequency bands, with linear fits. Zero (dashed black line) represents no mean change from rest (overlapping rest and move clusters). Shading represents a 95% confidence interval of the slope. The small gray upward arrow indicates where P2 transitioned between using both ankles to right hand as their primary control gesture. For visualization, Euclidean distances greater than 25 were clipped (dashed gray line), with the original value displayed above the corresponding scatter point. Scatter points with an outline are significantly different from zero (permutation test, p < 0.05). Scatter points with a black ring were identified as high influence points and not included in the linear regression. A total of 10 samples were flagged as high leverage, corresponding to a mean of 1.1 samples per participant × band. Linear fits with solid lines have non-zero slope (Wald test, p < 0.05). n.s., not significant and sig, significant.

With a few exceptions, most participants and bands did not exhibit a significant change over time in the Euclidean distance motor signal modulation metric. All participants reported here had ALS, a progressive neurodegenerative condition that may have contributed to observed decreases in attempted movement modulation, regardless of statistical significance. There was a significant decrease in the low gamma band for P2 with both ankles (p<0.01), however, there were no significant changes for any of the bands for P2 after they switched to the right hand. Additionally, P1 and P6 showed a significant increase in the beta band (p<0.05). These increases were accompanied by decreases in false positive rates and latency, and such changes may reflect learning and task familiarity.

### Relationship between online performance and neural modulation strength

Performance in the online command-control task was consistently high across participants (Fig. 4). For three participants, accuracy showed a modest increase over the follow-up (up to 6.0%/year), although the change was not statistically significant for any participant.

**Fig. 4:**
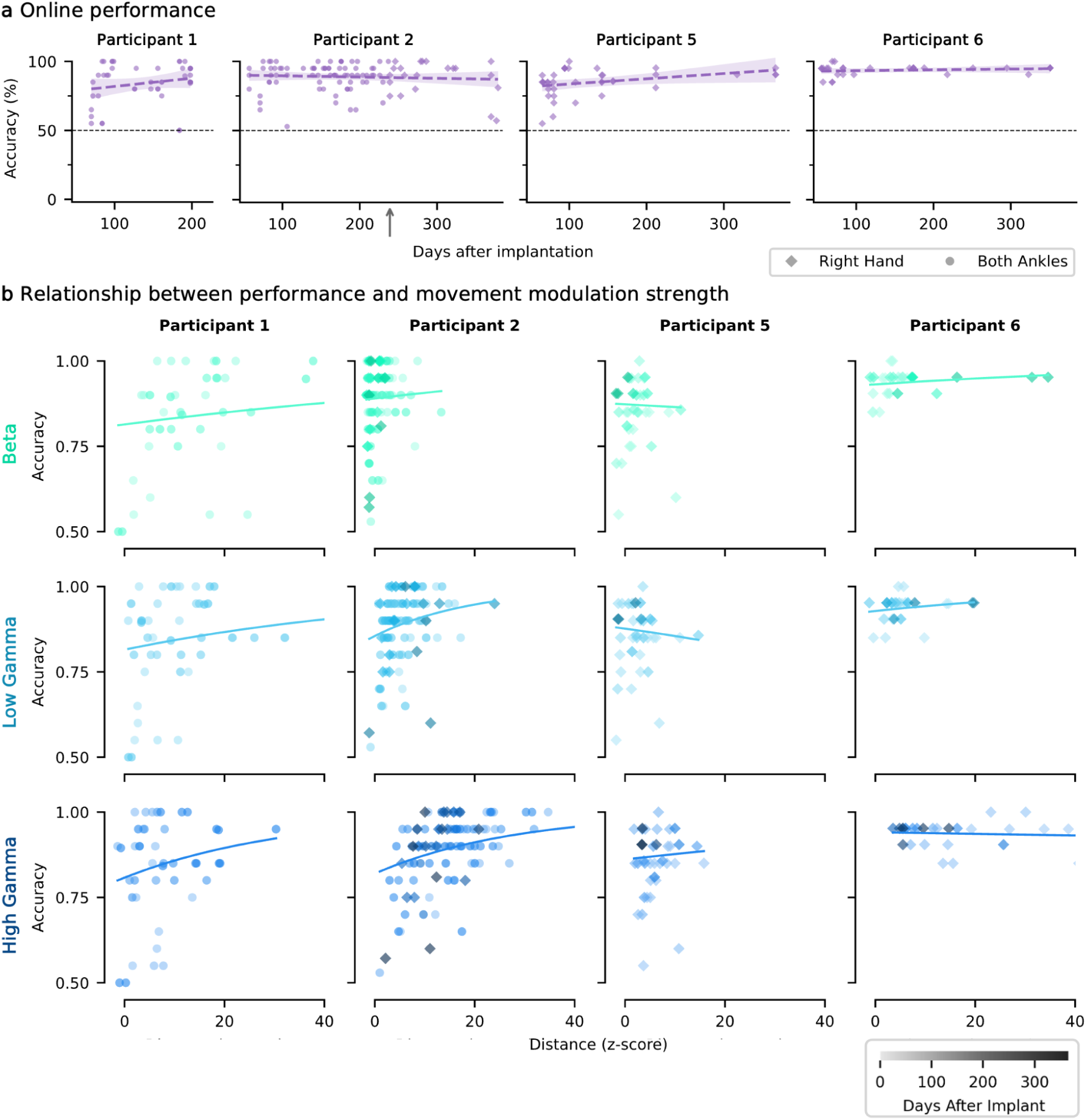
Relationship between online performance and neural modulation strength. **a**, Online accuracy over time with linear fits. Linear fits with solid lines have a significant slope (Wald test, p < 0.05). n.s., not significant and sig, significant. Shading represents a 95% confidence interval of the slope. Right hand movements are represented by diamonds, and both ankles by circles. The small gray upward arrow indicates where P2 transitioned between using both ankles to right hand as their primary control gesture. **b**, Online performance (accuracy) is plotted against movement modulation strength, measured as the cross-validated Euclidean distance between rest and move clusters, which was measured for each frequency band. The relationship was modeled with a saturating exponential function (1 − *ae^bx^*). Each scatter point represents a single session, with darker points indicating longer time post implant. The x-axis was limited to 40 for visualization purposes.

Next, we examined the relationship between online accuracy and modulation strength quantified with Euclidean distance. Across participants and bands, generally the accuracy increased as modulation strength increased.

### Resting state spectral features are stable over one year

To assess the stability of resting state signal features over time, 2-minute resting baseline recordings were made at the beginning of each home session. The mean band power in six canonical frequency bands was calculated for all channels. Examples of broadband resting state signals for a single channel and their corresponding PSD for P2 is shown in Fig. 5a. To assess changes in baseline band power over time, each channel (for all participants) was fitted with a linear regression to quantify the slope and determine its statistical significance. The number of channels exhibiting significant slope varied across frequency bands (range 4–52%), with higher-frequency bands generally showing a greater proportion of channels with no significant change over time (Fig. 5c). The overall distribution of slopes for all bands was centered or near zero, with higher frequency bands exhibiting tighter distributions around zero (Fig. 5c).

**Fig. 5:**
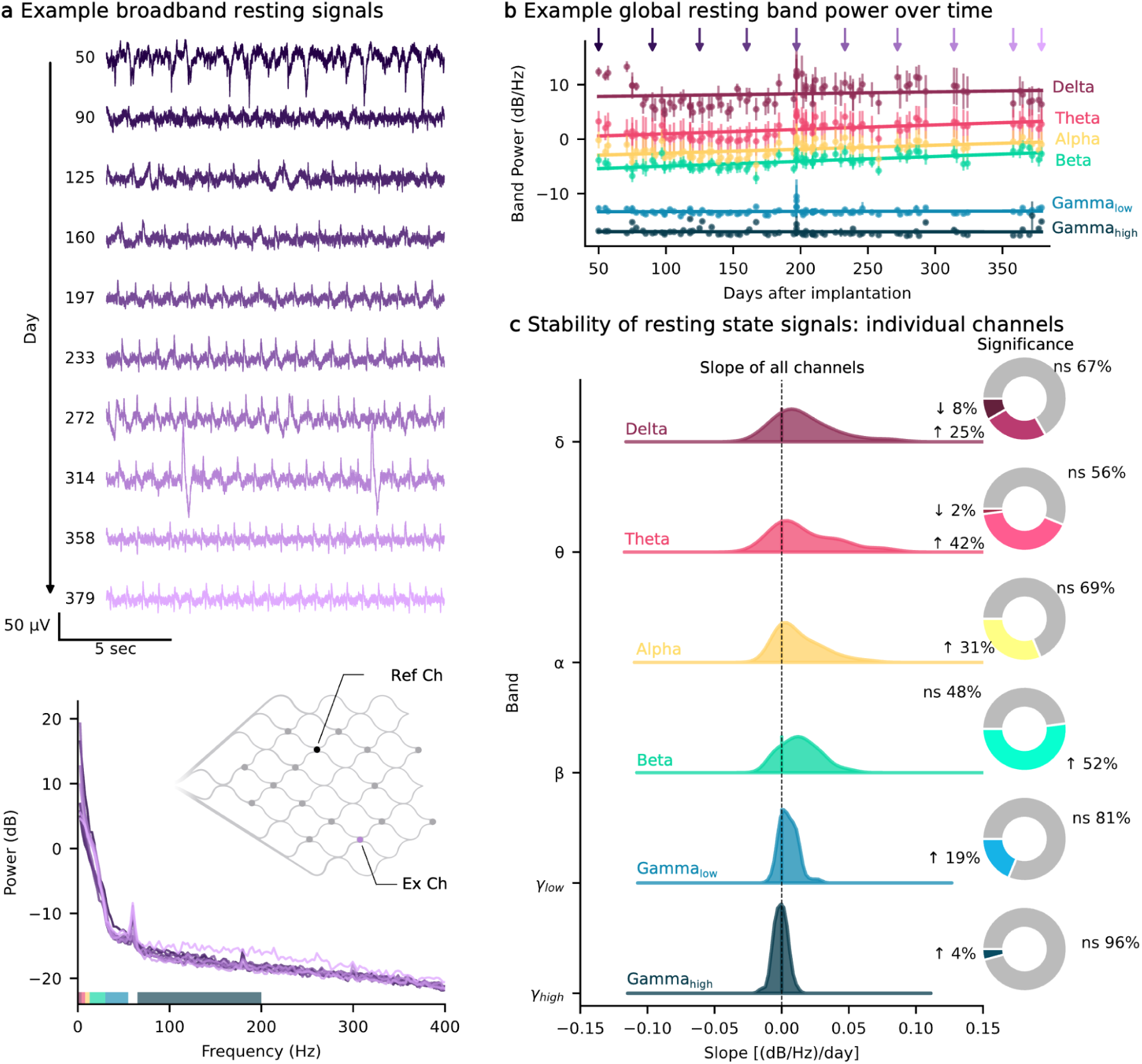
Stability of resting state signal features over time. **a**, Short recordings of broadband signals (2 min) were acquired while the participant was resting at the start of every training session. Top: Example resting-state broadband (high-passed at 1 Hz) signals for a selected electrode over the course of a year (from participant 2); bottom: Welch’s power spectral density (PSD) estimates of the corresponding resting state recordings. Diagram of example electrode location on the stent array. **b**, Mean (± standard error of the mean) band power across channels over the course of a year with linear fits to each frequency band (example from participant 2). The purple arrows correspond to the days shown in panel a. **c**, The band power over time for all channels across participants (P1, P2, P3, P5, P6) were modeled individually with linear fits. The distribution of slopes are estimated with a gaussian kernel density estimate. The pie charts show the number of channels that are significantly changing over time (Wald test, p < 0.05) and if they are, if the slope is positive (increasing) or negative (decreasing) as indicated by arrows. ns, non-significant.

In general, band power remained stable over time, with a large proportion of channels exhibiting no change in band power over the year following implant. Of those that have a significant slope, the average slope ranged (depending on frequency band) from 0.007 to 0.042 (dB/Hz)/day for increasing channels and -0.0035 to -0.012 (dB/Hz)/day for decreasing channels, representing small changes per day.

### Electrode impedances are stable throughout the testing period

To monitor the electrode-tissue interface over time, in-situ impedance measurements were recorded at the start of each home session. Across all participants, 11 of 64 electrodes were excluded by visual inspection or due to high impedance (>100 kΩ), leaving on average 10.6 active channels per participant (Extended Data Table 1). The mean impedance across all channels and all participants was 30.7 ± 15.8 kΩ, with a mean difference of 12.1 ± 12.7 kΩ from the reference electrode (Fig. 6, Extended Data Table 1). Among electrodes with impedances below 100 kΩ, 45% remained stable over time (Wald test, p>0.05), while 25% exhibited a significant increase and 30% showed a significant decrease. Among electrodes with increasing or decreasing impedance, the mean slope was 0.049 and -0.020 kΩ/day, respectively. Thus, even for electrodes that exhibited changes over time, the overall rate of change remained low. These stable impedance recordings over time suggest consistent electrode-tissue contact, minimal tissue response, and a stable recording environment, indicating long-term reliability of the implant.

**Fig. 6:**
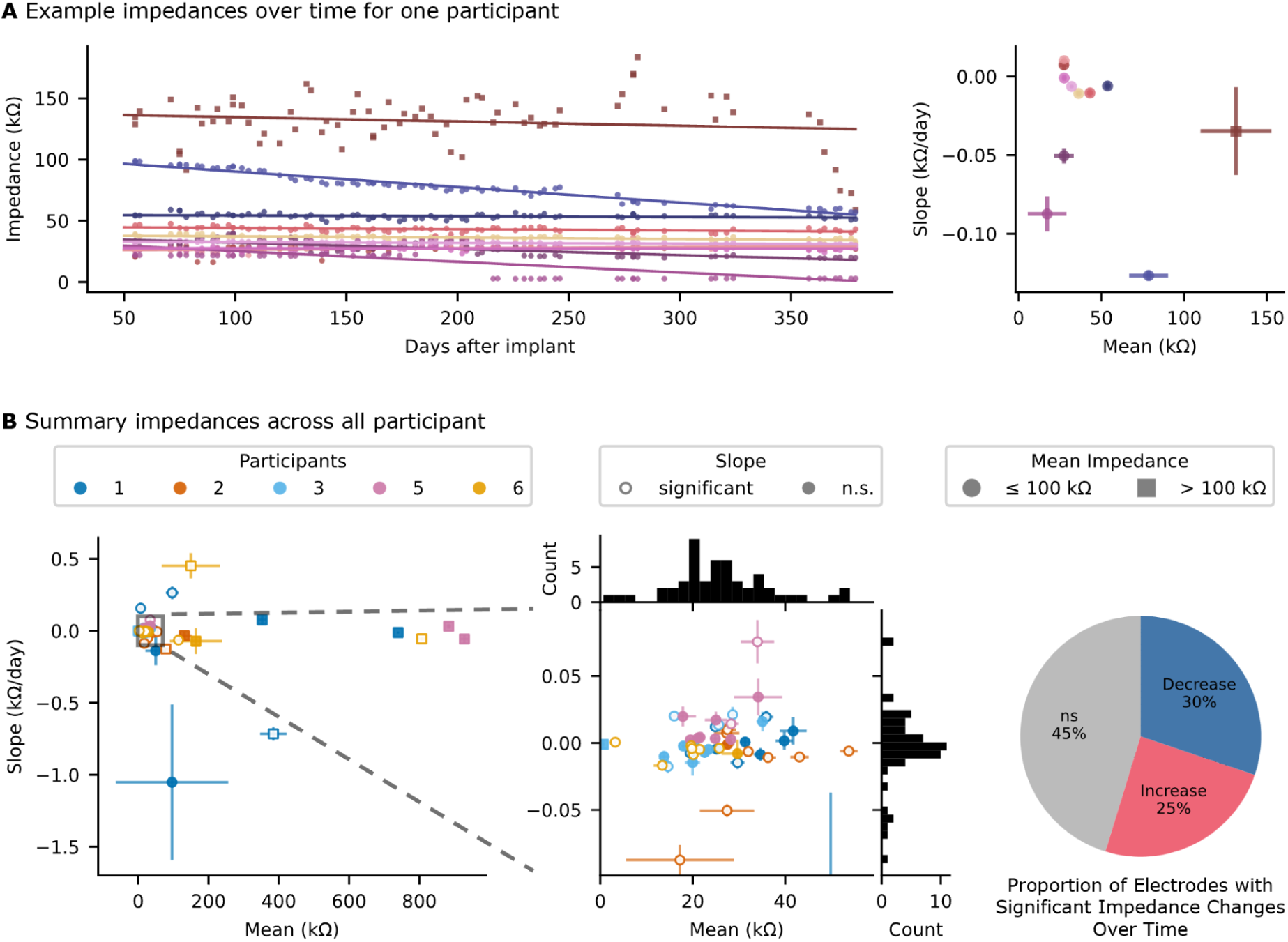
Impedances over time. **a**, On the left, example impedances for all electrodes plotted against days since implantation (for P2), with each electrode represented by color. Solid lines show the fit linear regression to each electrode. On the right, each electrode is summarized by a scatter point (with error bars) showing the mean impedance (std deviation) and slope (standard error of the estimated slope) from the linear regression fit. **b**, All electrodes for across the 5 participants (indicated by color for P1, P2, P3, P5, and P6) showing the mean (std deviation) and slope (standard error of the estimated slope) and zoomed in section showing the large cluster around 0 Ω/day and 20 to 40 kΩ (middle). Filled markers indicated electrodes with insignificant change in impedance over time (Wald test, p < 0.05) while unfilled markers indicated significant change. Electrodes with high impedance (mean greater than 100 kΩ) are marked with squares, the remaining electrodes are marked with circles. Pie chart illustrating the distribution of electrodes based on the significance and sign of the slope from the linear regression fit of impedances over time (for electrodes with low impedance). n.s. non-significant.

### Exploratory analysis of gesture discriminability for multi-switch control

Clear visual modulation of high gamma was observed during various cued attempted movements, relative to rest. Representative single trial and trial-averaged high-gamma envelopes are shown for P1 (Fig. 7a). While the magnitude of the responses varied across electrodes, the mean temporal profiles were similar across electrodes and gestures (Fig. 7a).

**Fig. 7:**
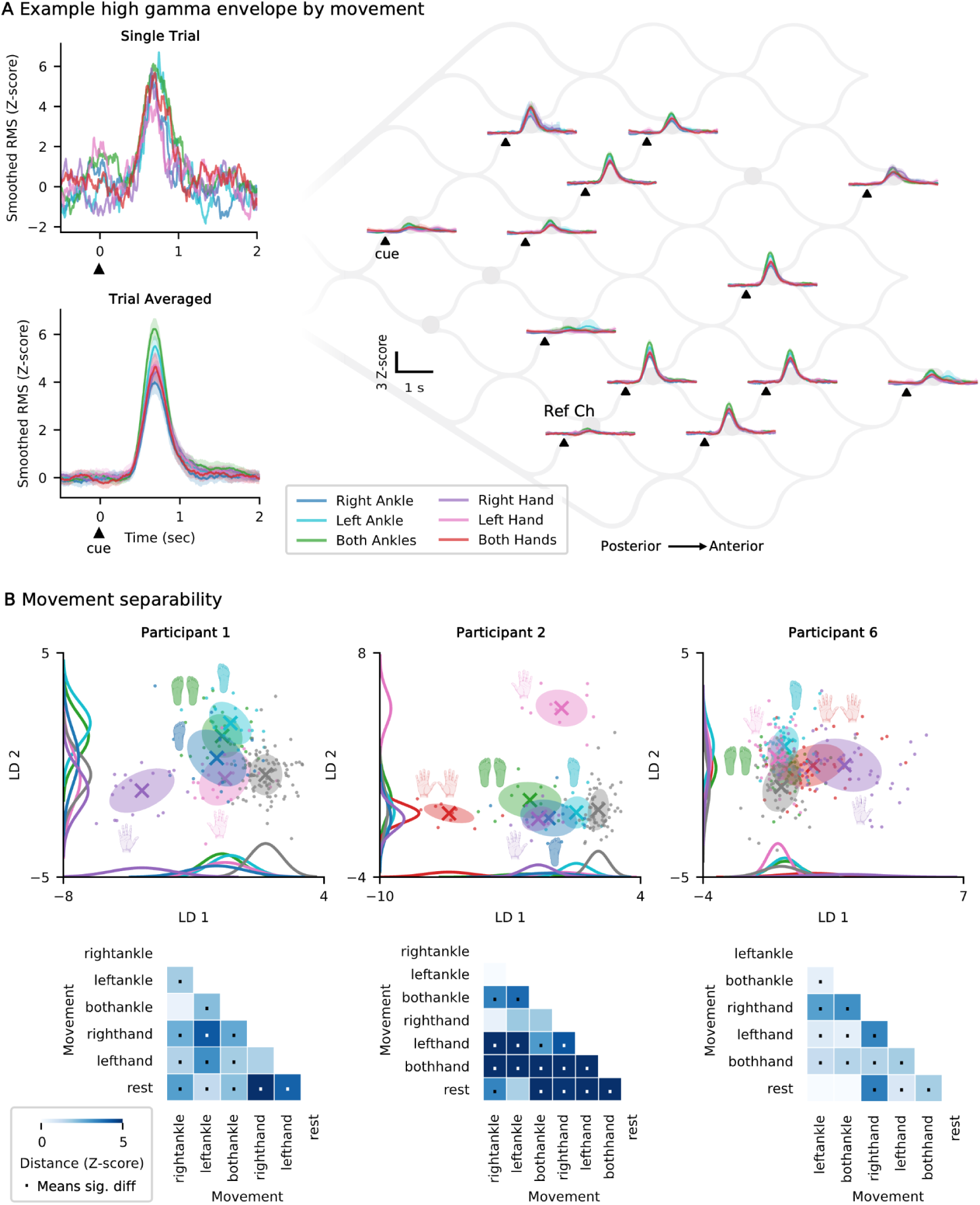
Exploration of multi-gesture discriminability. **a**, Top: Examples of smoothed high gamma envelopes when cued to perform a specific gesture, showing representative single trials. Each gesture is represented by a different color. Bottom: Each line shows the mean envelope across many attempts of the same gesture with shaded regions showing the 95% confidence interval of the mean (example shown for P1). Right: Same as the last panel but showing all active channels laid out spatially on the stent array. **b**, *Top*: The feature vector including beta, low gamma, and high gamma RMS for each movement projected into 2D space with LDA. Shaded ellipse shows the covariance and X marks the centroid. The distribution for each class is visualized using gaussian kernel density estimates along both dimensions. *Bottom*: The cross-validated Euclidean distance between each possible movement pair. Distances exceeding 5 were capped to improve visualization. A dot in the middle indicates the 95% confidence interval does not include 0, indicating the distance between cluster means is significantly greater than 0. RMS, root mean square; LDA, linear discriminant analysis

With the goal of identifying a subset of gestures that could be used for multi-class decoding (ex: two distinct switches), we analyzed data from a battery of gestures collected offline without feedback. Such multi-switch control supports functions such as left and right mouse clicks, which enables faster and more complex control of digital devices. Gesture discriminability was visualized using LDA with features pooled from beta, low gamma, and high gamma band power. The first two LDA dimensions, illustrating gesture-specific clusters are shown for each participant (Fig. 7b). The distance between clusters was quantified with cross-validated Euclidean distance (Fig. 7b). This exploratory analysis aimed to identify a subset of gestures that were both well separated from rest and from each other. For example, in P1, the right hand and left ankle signals show separability and may be a viable candidate for multi-click control. While these preliminary results are promising, further validation including real-time multi-gesture decoding is necessary to confirm these results. Furthermore, due to limited multi-class data, this exploratory analysis was restricted to three participants (P1, P2, and P6). This exploratory analysis on gesture discriminability was performed offline without feedback and thus is likely a conservative minimum for separability. With real-time neuro-feedback training and repeated practice, the neural responses may strengthen and become more distinct.

### Referencing scheme influences artifact presence

Data was obtained using one of two hardware reference schemes: (1) all channels referenced to a single selected electrode on the body of the stent or (2) all channels referenced to an electrode in the chest on the body of the IRTU in the subcutaneous pocket (Extended Data Fig. 1 a & b). When using the stent as a reference, one of the active electrodes was effectively lost due to the referencing scheme. However, this approach significantly reduced common-mode noise, particularly electrocardiogram (ECG) artifacts. Despite this reduction, residual ECG artifacts remained in some electrodes, as indicated by regularly occurring R peaks or T waves (Extended Data Fig. 1e). On average, across participants, 3.2 channels exhibited moderate or greater residual ECG artifacts. Importantly, ECG contamination was generally low frequency, predominantly below 40 Hz^23^, and did not substantially affect higher-frequency signals, particularly gamma, which were those used for decoding. Consequently, given the use of the local reference, it was not a major concern and was not further addressed. To visually demonstrate the reduction of the ECG artifact, the stent reference can be approximated by software re-referencing the IRTU referenced data through subtraction of a single channel (Extended Data Fig. 1c). For all reported online control results, the on-stent local reference was used. For baseline analyses, signals were also obtained using the local reference; in the rare case where an IRTU reference was used, the data was software re-referenced to a local channel.

Occasionally, other artifacts were noted, such as line noise which was easily identified by narrow peaks in the PSD at 60, 120, and 180 Hz (Extended Data Fig. 1e). Moderate line noise contamination, defined as an average peak prominence exceeding 5 dB, was observed in 31.3% (±5.3%) of sessions across participants. Notably, line noise contamination affected all channels similarly, with sessions either showing uniformly high or low 60 Hz prominence across channels. Since recordings were conducted in the home environment, we suspect that session-to-session variability in line noise was driven by changes in the surrounding electronics, such as televisions, space heaters, eye trackers, or variations in proximity to power outlets. Frequency bands were defined to exclude 60 Hz, but no further filtering or removal methods were applied as line noise was generally minimal.

## Discussion

Here we report on signal characteristics and present evidence of long-term stability of neural signals recorded endovascularly for motor-based iBCI control. Data was collected from participants with severe paralysis enrolled in a clinical trial evaluating an endovascular-based iBCI. We report signal characteristics over 1 year for three participants and 6 months for two participants. vECoG signal stability was assessed with a variety of metrics, including motor signal modulation, resting state signal features, and impedances. All data was collected in the participants’ homes, where real-world noise and disturbances pose greater challenges than a controlled laboratory setting, highlighting the robustness of the recorded signal under natural conditions. This study demonstrates the potential for endovascular iBCIs to reliably detect stable neural signals, which is necessary to enable at-home iBCI driven digital device control.

### Long-term stability of implanted brain-computer interfaces

Users of implanted BCIs should expect the devices to operate reliably for as long as they need them, perhaps several years. Aside from device related failures, a variety of biological factors may diminish BCI performance, including tissue reactions and neuronal loss. Preclinical studies have shown that endovascular arrays and leads become integrated into the vessel through endothelial encapsulation, with the struts moving deeper into the vessel wall over the first few days to weeks^24, 25^. Prior stenting literature reports that full endothelization occurs between 4 to 6.5 weeks^20,26^. Over this period, prior work demonstrates improved recording sensitivity, evidenced by better detectability of somatosensory evoked potentials^24^. In contrast, subdural ECoG arrays become encapsulated in fibrotic connective tissue following implantation,^27^ which can increase the distance between the cortex and the recording electrodes, potentially attenuating signal amplitude^28–30^. In either case, whether fibrous encapsulation or endothelialization, it is expected that the first few days following implantation will exhibit the most rapid changes in the electrode-interface and recorded signals. However, in this study, data collection began 6 to 8 weeks after implant to allow resolution of local bruising and swelling at the site of the subcutaneous IRTU. Therefore, we cannot assess the signal quality changes, but other neural interfaces have shown rapid changes in the acute postoperative period^24,30,31^. In chronic human BCI studies, ECoG electrodes have shown fluctuations in impedance over the first 1 to 6 months post implantation before stabilizing^30,32–34^. In this study, we did not observe large fluctuations in impedance from 2 to 12 months post implant. Overall, the largest proportion (45%) of electrodes did not change impedance significantly, and of those that did, the median change was -0.03% to 0.05% per day from the channel’s mean impedance. These small changes are consistent with other reports on in-situ impedance changes after implant^32,33,35^.

RMS voltage or band power during rest are commonly used metrics to assess long-term signal stability. There is growing evidence to support that ECoG-based recordings exhibit stable band power in chronic implants in humans^32,33,36,37^. In our study across 1 year, we found a mix of changes in power across all bands, with a large proportion of channels unchanged, some channels increasing, and some decreasing. However, even when changes were detected as statistically significant, the actual changes were relatively small, ranging from -0.012 to 0.042 (dB/Hz)/day. For comparison, previous studies have reported modest and variable changes in band power over time, including early postoperative changes followed by small increases or decreases depending on the individual and study^32,36–38^. Overall, the power changes observed in this study are consistent with prior reports on chronic ECoG implants in humans, which similarly describe only minor fluctuations over time. Overall, the long-term recordings in this study demonstrate that stent-electrode arrays can record stable signals for at least 1 year. Importantly for long-term control, the stable rest state recordings suggest no significant underlying changes in the population neural dynamics that would necessitate recalibration or other intervention.

### Detecting motor intent: gamma synchronization

Consistent with earlier literature, gamma and high gamma signals showed clear synchronization during attempted movement. Several ECoG studies have shown increased gamma-band power during executed movements, attempted movements, and motor imagery^12,15,16,19,35,39–44^ and has supported many BCI applications including decoding of movement trajectories ^39,40^, movement onset and offset^44^, and multiple independent control signals^35,40^. Here, we observe strong movement related gamma synchronization, reaching on average 10.9 standard deviations above rest, demonstrating that high frequency motor signals can be recorded from within the vasculature. Despite this unique location and electrode geometry–nestled in the endothelium and surrounded by dura–we found that the motor-related features of the vECoG signals are similar to those reported for subdural ECoG grids placed on the cortical surface. By recording intracranially through the vascular system, this approach bypasses the filtering effects of the skull and scalp, enabling the reliable measurement of high-frequency features that are attenuated and difficult to resolve without trial averaging on scalp electroencephalography (EEG) recordings.

Furthermore, our results are consistent with previous observations of retained gamma-band modulation in people with ALS, including those with advanced paralysis^19,45–47^. The reliable decoding of gamma synchronization with this sample supports the potential utility of vECoG BCIs as a motor neuroprosthesis for long-term communication and digital device control.

One of the primary advantages of gamma band signals is their spatial specificity. These spatially specific activation maps have demonstrated successful decoding of multiple degrees of freedom, such as decoding multiple complex hand gestures,^35,48,49^ bimanual exoskeleton control,^50^ and two-dimensional cursor control^40^. Here, we analyzed six gestures to evaluate the potential for multiple distinct switches and found a subset of gestures that were separable, which is consistent with prior studies reporting successful classification of multiple movement types using high-frequency spectral features^35,49,51^. Despite methodological differences, our findings support the broader conclusion that gesture-specific neural activity is distinguishable with vECoG. By prioritizing a small set of gestures for multi-switch control rather than maximizing the number of classes, this approach addresses the clinical need for robust control in users with severe disability. In comparison, the information captured with vECoG is likely less complex than that recorded with intracortical MEAs, which have enabled fine-grained motor decoding with high accuracy, such as tracking individual finger trajectories^9,52^. In contrast, the vECoG records population-level field potentials, which may limit resolution but affords long-term stability. Nonetheless, these results suggest that two or more gestures could serve as discrete switches for clinical vECoG BCI applications, such as assistive device control.

In many participants, the gestures identified for potential multi-switch control included hand movement. Given the position of the stent-electrode array within the superior sagittal sinus near the paracentral lobule, which classically controls lower limb movement, it is anticipated that motor attempts involving the lower limbs would generate the strongest signals. However, we found strong representations of both ankle and hand movement (Fig. 7b). This may reflect factors such as proximity to the supplementary motor area (SMA) or mixed effector regions throughout motor cortex^53,54^.

### Detecting motor intent: beta desynchronization

Beta desynchronization, typically observed as a reduction in beta-band power (13–30 Hz) during movement or motor imagery, is a well-established neural correlate of motor intention^13,42^. In our cohort of participants, we reliably saw movement related gamma synchronization in 4 out of 5 participants, while beta modulation was seen only in 2 out of 5 participants. This may have been due to 1) close referencing channel and/or 2) ALS disease progression. Firstly, the detection of beta desynchronization was more sensitive to the type of referencing scheme, which included a remote reference in the chest or a local reference channel on the stent-electrode array. In this study, the local reference scheme was preferred, because it provided greater attenuation of common mode noise sources, especially electrocardiogram artifacts. However, the close proximity of the local reference and recording electrodes may have resulted in stronger attenuation of beta band signals, which tend to exhibit high degrees of correlation across electrodes spaced 2-4 mm apart^55^. On the other hand, difficulty detecting beta band modulation may also reflect ALS disease progression. A previous report on a person with locked-in syndrome due to ALS demonstrated variable and weak decreases in low-frequency band activity during attempted movement, even in electrodes showing the highest gamma response^46^. Scalp EEG studies have similarly reported that ALS patients have significantly smaller magnitude event related desynchronization during motor imagery than age-matched controls^56^. Furthermore, in ALS patients, the severity of bulbar impairment was correlated with smaller event related desynchronization^56^. Further, the majority of our participants were non-verbal, ventilator dependent, and reliant on percutaneous endoscopic gastrostomy (PEG) for nutrition, indicating severe bulbar impairment, which may explain the weak beta modulation. These findings suggest that while beta desynchronization is a commonly reported response during motor attempt, it is not universally present in all individuals or under all recording schemes.

### Study limitations

Although the initial follow-up reported here is 6-12 months, it is imperative to conduct ongoing assessments over several years to confirm neural signal stability and reliability. There are open questions about how ALS related cortical degeneration of motor neurons affects BCI device usability^45^. Additionally, all reported participants had ALS, therefore more data is needed to assess if there are etiology-specific differences.

Here we present data on the characteristics and longevity of signals recorded from endovascular stent-electrode arrays, which is one of many factors that will shape usability and long-term success of iBCIs. A significant limitation of this paper is the absence of clinical outcomes, including adverse events, which will be addressed in other publications. Specifically, ensuring that the rates of device migration, thrombosis, intracranial hemorrhage, stenosis, and infection are acceptably low for the provided benefit remains essential.

## Conclusion

The data presented herein demonstrates stable neural signals recorded with an endovascular BCI in three participants over a year and two participants over six months of follow up. This long-term stability is a critical foundation for enabling reliable, independent, at-home digital device control. As the technology continues to evolve, ongoing evaluation of the signal properties and longevity will be essential to support scientific understanding.

## Methods

### Clinical Trial and Device

The data presented here were recorded as part of an early feasibility clinical trial to obtain preliminary evidence of the safety and efficacy of an endovascular BCI for digital device control and communication in people with severe paralysis (ClinicalTrials.gov: NCT05035823, WCG IRB: 1347924, Investigational Device Exemption: G210178). For all participants, severe upper limb motor impairment or paralysis prevented physical interaction with a personal computer, leading to either complete non-use or reliance on adaptive or augmented control methods.

The deployment of the stent-electrode array into the superior sagittal sinus via a catheter guided by angiography has been described previously^20,21^. The stent-electrode array (Stentrode, Synchron, NY, USA) consists of sixteen platinum electrodes embedded on a nitinol stent scaffold connected to a flexible lead, which is tunneled to an internal receiver telemetry unit (IRTU) placed in a subcutaneous pocket below the clavicle (Fig. 1). Data was transmitted to an external computer by placing an external receiver telemetry unit (ERTU) on the skin, allowing wireless infrared communication to the IRTU. Biopotential measurements on each electrode were referenced to a common electrode, which was located either at the IRTU or 1 of the 16 electrodes on the stent-electrode array. The sampling rate was 2,000 Hz per channel.

### Data Collection Sessions

User training and testing with the BCI began after all bruising around the chest pocket had resolved and infrared communication could be established, approximately 6-8 weeks after implantation (mean 7.2 ± 1.2 weeks). In-home sessions occurred approximately twice weekly, typically lasting 2-3 hours or as long as the participant felt comfortable. Sessions were not completed during some weeks due to personal breaks (e.g. holidays), health-related issues unrelated to the study, or wound healing. This paper analyzes session data up to 12 months post-implant, with two participants (P1 and P3) having shorter follow-ups.

Sessions were conducted at the participants home with a field engineer present. The engineer completed the system setup, which consisted of placing the ERTU over the skin above the IRTU and turning on the computer, and to explain new tasks and answer any questions. None of the participants had previous BCI experience. Tasks progressed in difficulty as the participant demonstrated proficiency, although here we only examine a subset of tasks to evaluate signal properties and stability.

### Baseline Evaluation

A baseline recording was taken at the beginning of every training session. The baseline task consisted of a two-minute recording with the participant resting with their eyes open and gazing at a fixation cross or circle on the computer screen in front of them. These recordings were utilized to analyze the resting state signal characteristics over time. In the few cases where the IRTU reference was used, the data was re-referenced in software to the participant’s standard local reference channel before further analysis.

Additionally, at the beginning of every home training session, impedance measurements were taken by applying a sinusoidal pulse (100 Hz, 10 nA) to each electrode. Three short test pulses were applied and the average of the three was considered to be the impedance value. Only the magnitude and not the phase was recorded.

### Pre-processing - Channel Selection and Artifact Handling

Any unconnected channels (mean 3.2 channels unconnected, see Extended Data Table 1) were removed prior to analysis. Furthermore, electrodes with high impedance (> 100 kΩ) or appeared noisy on visual inspection were excluded from further analysis (mean 2.2 channels excluded, see Extended Data Table 1), the remaining channels were designated as active channels.

Artifacts were detected during offline data analysis if they exceeded eight standard deviations from the mean, with a 50 ms buffer applied to each side, using thresholds derived from the mean and standard deviation of the initial ten baseline recordings. This removed on average 0.33% of the baseline data (P1: 0.41%, P2: 0.24%, P3: 0.09%, P5: 0.32%, P6: 0.61%).

To assess the extent of line noise, sessions were evaluated based on the prominence of the 60 Hz peak in the power spectral density (PSD). Prominence was calculated for each channel and run by computing the difference between the peak power within the 59–61 Hz range and the average power across the adjacent frequency bands (50–58 Hz and 62–70 Hz). The resulting prominence values were then averaged across all channels, excluding the reference channel. To assess line noise prevalence, the percentage of baseline runs in which the average prominence exceeded 5 dB was computed.

The presence of residual ECG was assessed by visually identifying channels with moderate residual ECG in baseline (resting state) recordings at 3, 6, and 12 months. A channel was marked as having residual ECG present if regularly occurring R or T waves were visually identified as being substantially larger than the background neural activity.

### Offline Analysis of BCI Performance - Motor Mapping Task

In the early testing sessions, participants undertook a battery of attempted movement training tasks, termed motor mapping tasks. These tasks were visually prompted on a computer screen and consisted of 10 trials of 5-s (± 1 s) rest periods where the participants focused on a fixation circle, followed by a 5-s period of movement attempt, in which 5 repetitions (1 Hz) of attempted movement occurred. The movements attempted included: both ankles, right ankle, left ankle, right hand, left hand, and both hands.

### Offline Analysis Metrics

Signal features were assessed in the standard frequency bands: delta (1-4 Hz), theta (4-8 Hz), alpha (8-13 Hz), beta (13 to 30 Hz), low gamma (30 - 55 Hz), and high gamma (65 - 200 Hz)^57,58^. Filters were implemented with second order zero-lag Butterworth filters. The power spectral density was approximated with Welch’s method. Bandpower was calculated for each frequency band, defined in the frequency band from [f_1_, *f*_2_] as^36^:

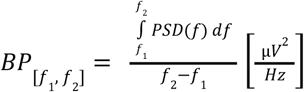

Spectrograms and band-limited signal envelopes were used throughout to visualize the data. The spectrograms were calculated using signal.spectrogram (SciPy library). To facilitate the identification of task-related changes in the spectrograms, the power values were z-scored by normalizing them to the mean and standard deviation of the corresponding frequency bins during the rest period. Signal envelopes were extracted by calculating the analytic amplitude of the Hilbert-transformed band-limited signals. These envelopes were then smoothed using either a 100 ms or 250 ms moving average and further z-scored based on the mean and standard deviation of all rest periods on a given day.

To assess the strength of the movement modulation between rest and attempted movement, we quantified the distance between the band power across conditions. Distances between distributions of band power vectors (ex: rest and right ankle movement) were measured using cross-validated Euclidean distance as described in Willet et al 2020^53^. This was done following the leave-one-out if the distributions had equal number of observations (ex: rest vs single movement) and splitting the data into folds when there were unequal observations (ex: rest compared to multiple gestures) where the number of folds were equal to the number of observations of the smallest class^53^. Pre-processing consisted of computing band-limited signal envelopes and smoothing with 100 ms moving average. Root mean square (RMS) vectors for rest and go epochs were computed and then z-scoring to the mean and standard deviation of the resting band power. For the rest period, RMS was calculated over the 200 milliseconds preceding the attempted movement cue onset. For the movement period, the RMS was computed from -100 to 100 milliseconds around the movement decode, or at the end of the movement period if no decode occurred. To determine if the Euclidean distance between the rest and move distributions was significant, a one-sided permutation test was used.

### Assessing Long-term Stability

To assess trends in features over time, linear regression was utilized to quantify changes in impedance, baseline band power, signal modulation, and online accuracy. The Wald test (Python SciPy) was used to determine whether regression slopes differed significantly from zero, thereby identifying channels/metrics with significant increases or decreases over time. Slopes that did not reach significance have insufficient evidence to conclude that they differ from zero. The specified metric was modeled as the dependent variable, while the days since implantation as the independent variable. To control for multiple comparisons, p-values were corrected using the Benjamini-Hochberg false discovery rate procedure. Corrections were applied separately within each participant across frequency bands and/or channels, depending on the analysis. Unless otherwise mentioned, p < 0.05 was used to determine significance.

For signal modulation measured with Euclidean distance, some data points exhibited high influence. This was particularly prominent when early or late sessions showed very high signal modulation, which produced substantial slope changes in leave one out diagnostics. To manage this, individual runs were assessed for high influence using Cook’s distance computed from ordinary least squares models fitted for each participant and band. Observations were flagged as influential when exceeding a threshold of 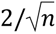, where n is total runs for a given participant. Flagged observations were excluded from the linear regression used to estimate the trend but still included in the scatter plots for visualization to preserve data context.

### Offline Multi-gesture Evaluation

To analyze differences in movements, offline motor mapping tasks across a variety of movements were analyzed. The attempted movements included: both ankles, right ankle, left ankle, right hand, left hand, and both hands. To minimize the risk of confounding factors related to session-specific variations, data from a single session with the highest number of gestures was analyzed. Since the comparison of multiple gestures was not a primary focus of the study, these runs were not conducted at regular intervals. To visualize the movement responses, the smoothed high gamma envelopes for various movements are shown.

To evaluate the feasibility of multi-switch control using vECoG, we analyzed the separability of neural features across multiple gestures. Band power was calculated for the rest period by taking the 1 second prior to the attempted movement cue onset, and for the movement period, by taking the 1 second after the movement cue, shifted by 500 ms to account for reaction time. P2 performed the gestures more slowly, so 2 seconds were used instead of 1. Beta, low gamma, and high gamma features were combined into a single feature vector. This vector was used for both visualizing the data in two dimensions by applying LDA (for visualization purposes, no cross-validation) and for applying cross-validated Euclidean distance to measure the distance between various gestures. To determine if the Euclidean distance between the two distributions (gestures) was significant, 95% confidence intervals were computed using jackknife resampling, performed once for each of the N trials (i.e. systematically leaving one trial out at a time). A distance was considered significant if the confidence interval did not include zero.

### Online Evaluation of BCI Performance

These online tests, termed command control test, were visually prompted on a computer screen and consisted of 10 trials of 10-s rest periods where the participants focused on a fixation circle, followed by a period of up to 10-s where the participant was visually cued to attempt a specific movement (“Move your right ankle”). When an attempted movement was decoded (also referred to as a “click”), auditory and visual feedback were provided: a ping sound was played, and a feedback bar filled. Additionally, the task automatically advanced to the next trial if a successful click occurred within the move cue period. At a minimum, these runs were completed at the 3-, 6-, and 12-month timepoints post-implant but were generally performed more frequently. This task was chosen for analysis because it was performed regularly and represented one of the most basic tasks, enabling straightforward characterization of signal properties and stability.

### Online Neural Decoders and Evaluation Metrics

The decoders utilized were generally based on real-time detection of transient oscillatory and pseudo-oscillatory neural activity^59^. A generalized algorithm used by most participants is described as follows. The neural data was first reduced to channels of interest based on prior evaluations of feature discriminability. The selected channels were then bandpass filtered into several gamma frequency bands. The resulting signals were then transformed into power-related metrics to quantify neural activity within each frequency band. To identify transient bursts, power and duration thresholds were applied (tuned for each participant and gesture), such that an event is classified as a burst when signal power exceeds the threshold for a minimum duration^59^. To determine if a switch occurred, a threshold on the number and rate of change of smoothed burst events was utilized. To prevent artifactual switches, if the feature was above a high threshold, the switch would be prevented. The system updated and produced predictions at 100 ms intervals.

Accuracy was measured as a percentage of trials classified correctly as either rest or move during the Command-Control Task. Specifically, a trial is deemed a successful rest (true negative) if no clicks were decoded during the cued rest period, while a trial is classified as a successful move (true positive) if a click was accurately decoded during the cued go period.

The decoder generated predictions every 100 milliseconds, providing a continuous stream of classification outputs throughout each trial. However, accuracy is evaluated based on the overall success of each trial, considering whether the trial was correctly classified as rest or move, rather than evaluating the accuracy of individual predictions within the trial. This approach emphasizes the system’s effectiveness in achieving the intended outcome for each trial as a whole.

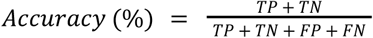

To examine the relationship between accuracy and signal modulation strength, classification accuracy was plotted against cross-validated Euclidean distance. The relationship was modeled using a saturating exponential function, 1 − *ae^bx^* where *a* scales the vertical offset and *b* the rate of increase. This model was chosen based on the expectation that accuracy increases approximately linearly with low modulation strengths but asymptotes near 1 (100%) as modulation increases.

## Data Availability

Source data for this study are available by reasonable request in the Data Archive for the Brain Initiative, with the identifier https://dabi.loni.usc.edu/dsi/1UG3NS120191. The analyzed datasets generated during the study are available for research purposes from the corresponding author on reasonable request once the full article is published in a peer-reviewed journal.

## Extended Data

**Extended Data Table 1:** Channel Information & Impedances.

| Participant ID | Data Follow-up<br>(months) | # Connected<br>channels* | # Active<br>channels† | Impedance<br>(mean ± std dev<br>kΩ) | Impedance<br>difference from<br>reference<br>(mean ± std dev<br>kΩ) |
| --- | --- | --- | --- | --- | --- |
| 1 | 6 | 16 | 13 | 41.0 ± 25.8 | 17.2 ± 19.9 |
| 2 | 12 | 11 | 9 | 32.5 ± 10.1 | 13.0 ± 6.7 |
| 3 | 6 | 13 | 12 | 22.7 ± 6.8 | 7.9 ± 4.3 |
| 5 | 12 | 12 | 10 | 25.0 ± 4.8 | 4.7 ± 3.5 |
| 6 | 12 | 12 | 9 | 32.3 ± 31.7 | 17.6 ± 29.0 |

\* Maximum channels on the stent-electrode array is 16.
† Includes channels that have in-situ impedance less than 100k $\Omega$ and visually clean.

**Extended Data Fig. 1:**
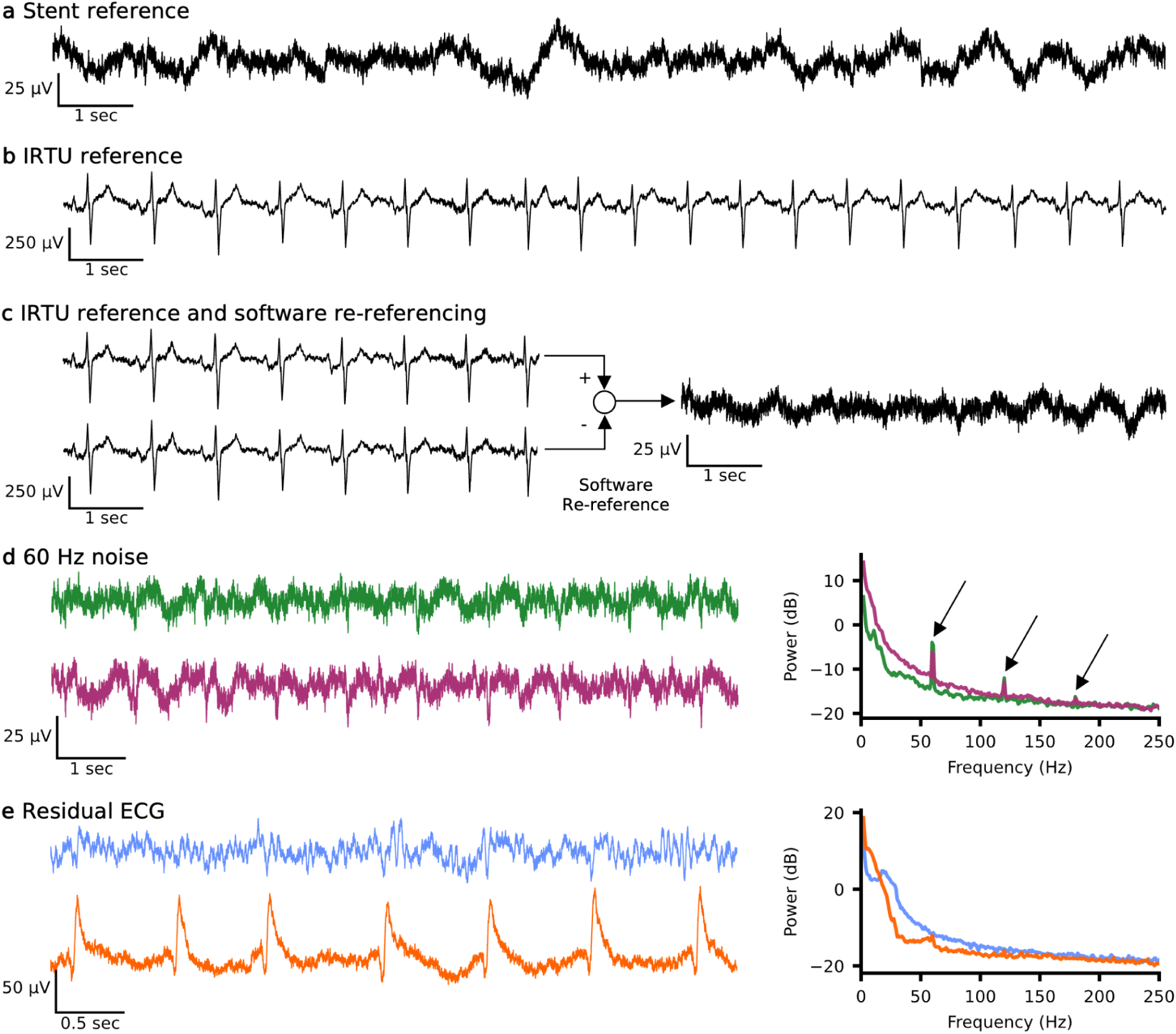
Referencing schemes and artifacts. **a**, Example resting data recorded with the reference channel selected from 1 of the 16 electrodes on the stent. **b**, Example resting data recorded with the reference channel selected to the IRTU in the subcutaneous subclavicular pocket exhibiting visual presence of ECG. **c**, Example resting data from 2 channels recorded with the reference channel selected to the IRTU (left) and the resulting signal when the data is software re-referenced to the bottom channel demonstrating a visual reduction in the ECG present. **d**, Example of a data recording where line noise was present on all channels. Example time series of two channels (left) and corresponding power spectral density using Welch’s method showing all channels (thin, colored lines) and mean of all channels (thick, black line) with arrows pointing to the frequency of line noise and its harmonics (60, 120, and 180 Hz). **e**, Example of variations in the residual ECG signals using hardware stent reference. The top shows minimal residual ECG, while the bottom displays moderate residual ECG, both recorded simultaneously during rest. IRTU, internal receiver transmitter unit; ECG, electrocardiography

## Acknowledgments

We thank all of our participants and their care-partners for their dedication to this study. We thank Marta Lapinska, Sara Onesi, Jacquelynn Jones, and Gianna Coscia for their invaluable assistance with participant recruitment and clinical coordination. We thank Ryley Bishop, Brian Franco, Maria Nardozzi (Tarbell), Ragan Chizmar for their support in data collection. We thank PJ Abels for her administrative and grant support. We thank Devapratim Sarma and Ashley Dalrymple for their early contributions to grant and protocol development.

## Author Contributions

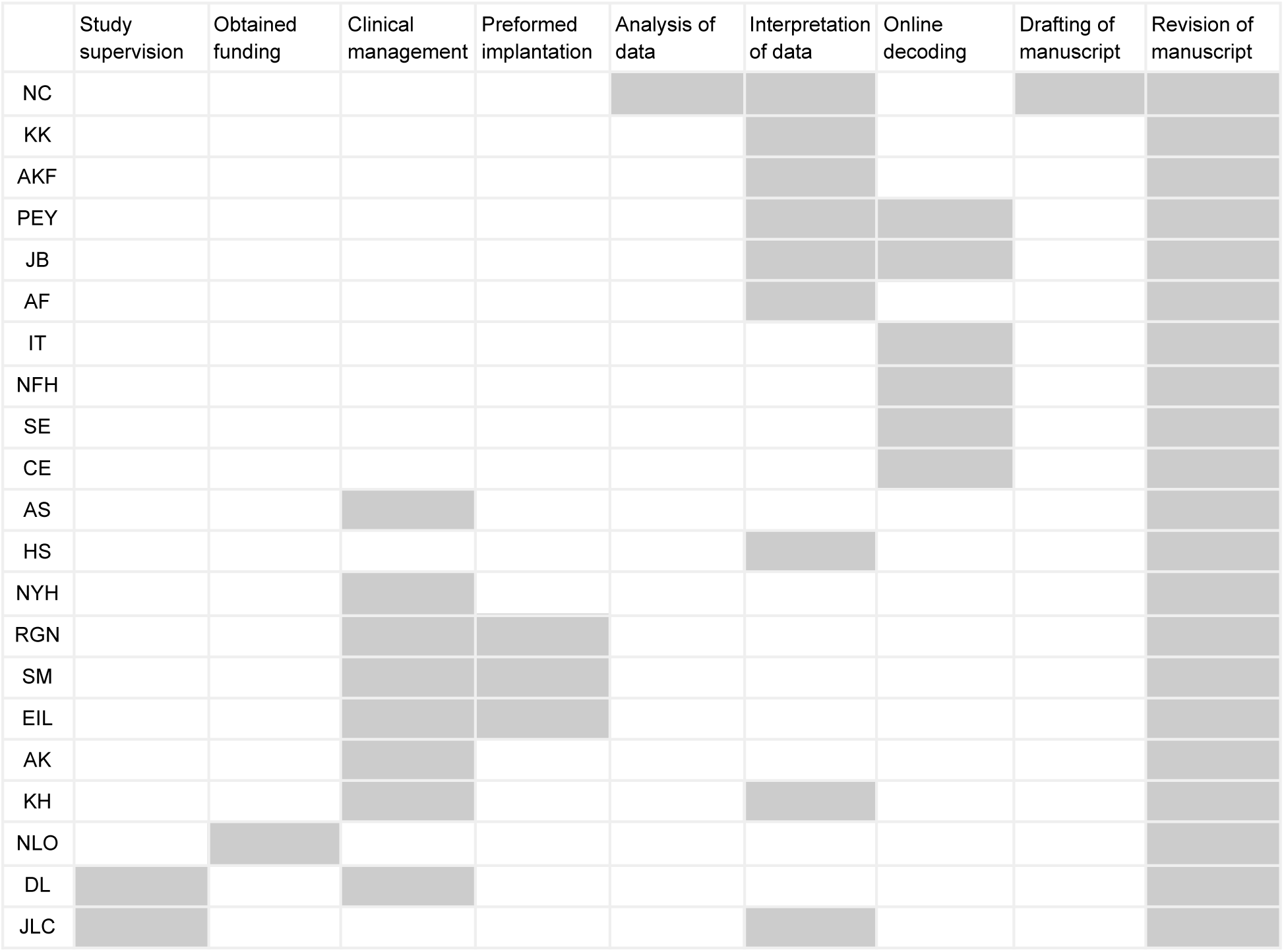
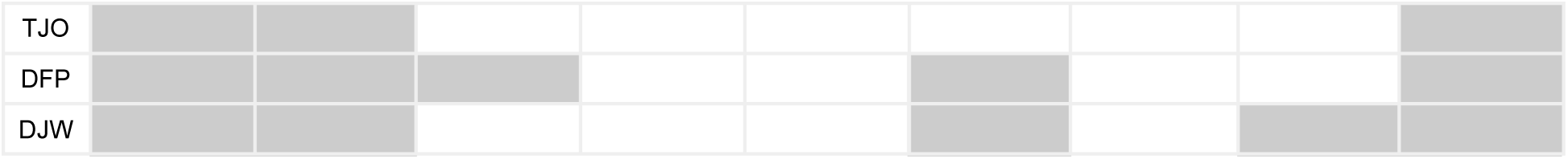

